# High Definition transcranial Direct Current Stimulation (HD-tDCS) for chronic tinnitus: outcomes from a prospective longitudinal large cohort study

**DOI:** 10.1101/2020.10.02.20173237

**Authors:** Laure Jacquemin, Griet Mertens, Giriraj Singh Shekhawat, Paul Van de Heyning, Olivier M. Vanderveken, Vedat Topsakal, Willem De Hertogh, Sarah Michiels, Jolien Beyers, Julie Moyaert, Vincent Van Rompaey, Annick Gilles

## Abstract

**Background:** Transcranial Direct Current Stimulation (tDCS) aims to induce cortical plasticity by modulating the activity of brain structures. The broad stimulation pattern, which is one of the main limitations of tDCS, can be overcome with the recently developed technique called High-Definition tDCS (HD-TDCS).

**Objective:** Investigation of the effect of HD-tDCS on tinnitus in a large patient cohort.

**Methods:** This prospective study included 117 patients with chronic, subjective, non-pulsatile tinnitus who received six sessions of anodal HD-tDCS of the right Dorsolateral Prefrontal Cortex (DLPFC). Therapy effects were assessed by use of a set of standardized tinnitus questionnaires filled out at the pre-therapy (T_pre_), post-therapy (T_3w_) and follow-up visit (T_10w_). Besides collecting the questionnaire data, the perceived effect (i.e. self-report) was also documented at T_10w_.

**Results:** The Tinnitus Functional Index (TFI) and Tinnitus Questionnaire (TQ) total scores improved significantly over time (p_TFI_ < .01; p_TQ_ < .01), with the following significant post-hoc comparisons: T_pre_ vs. T_10w_ (p_TFI_ < .05; p_TQ_ < .05) and T_3w_ vs. T_10w_ (p_TFI_ < .01 ; p_TQ_ < .01). The percentage of patients reporting an improvement of their tinnitus at T_10w_ was 47%. Further analysis revealed a significant effect of gender with female patients showing a larger improvement on the TFI and TQ (p_TFI_ < .01; p_TQ_ < .05).

**Conclusions:** The current study reported the effects of HD-tDCS in a large tinnitus population. HD-tDCS of the right DLPFC resulted in a significant improvement of the tinnitus perception, with a larger improvement for the female tinnitus patients.

## 1. Introduction

Subjective tinnitus is the perception of a sound in the absence of an internal or external source, which is often perceived as sizzling, hissing or ringing (Baguley et al., 2013). Approximately 10-15% of the adult population experiences this symptom, with 2.4% reporting a considerable amount of distress and a negative impact on their quality of life (Baguley et al., 2013, Axelsson and Ringdahl, 1989, Cardon et al., 2019). It is hypothesized that tinnitus results from maladaptive brain plasticity following hearing damage, which involves a wide network of cortical areas (e.g. central auditory system) and subcortical structures (e.g. limbic system). However, the precise pathophysiology of tinnitus is not yet fully understood (De Ridder et al., 2014, Baguley et al., 2013).

The cerebral cortex can be stimulated with transcranial Direct Current Stimulation (tDCS), which is a neuromodulation technique that aims to induce cortical plasticity and modulate the activity of the malfunctioning brain structures responsible for tinnitus (Langguth and De Ridder, 2011). The two frequently used sites of stimulation documented in literature for tinnitus relief are: Left Temporal Area (LTA) or Dorsolateral Prefrontal Cortex (DLPFC) (Song et al., 2012b, Fregni et al., 2006, Rabau et al., 2017, Shekhawat et al., 2013, Faber et al., 2012, Frank et al., 2012, Lefaucheur et al., 2017). There exists only limited evidence on the neurophysiological basis of tDCS due to the involvement of multiple brain areas in tinnitus and the non-focal stimulation by tDCS. It might be that tDCS of the LTA also activates surrounding cortical areas which, by inhibitory connections or neural competition, decreases the activity of other tinnitus-related areas (Fregni et al., 2006). Stimulation of the DLPFC, on the other hand, may strengthen deficient inhibitory top-down mechanisms in tinnitus, inducing auditory sensory gating in the anterior cingulate cortex (ACC) (Song et al., 2012b). Moreover, a resting-state electroencephalography (EEG) study by Vanneste and De Ridder (2011) showed that tDCS of the DLPFC was able to suppress tinnitus by modulating the primary auditory cortex, the parahippocampus, and the ACC. Hence, tDCS of the DLPFC influences the functionally connected tinnitus-related areas.

The effect of tDCS on the tinnitus perception was reviewed by Song et al. (2012b). They found that 39.5% of the tinnitus patients responded to tDCS with a mean tinnitus intensity reduction of 13.5% on a visual analogue scale for loudness (VAS). However, according to Lefaucheur et al. (2017), there is level of evidence B (i.e. probable inefficacy) for anodal tDCS of the LTA to relieve chronic tinnitus. In other words, the clinical effects of stimulating the LTA for tinnitus are probably absent. Recommendations regarding the potential efficacy of tDCS targeting the DLPFC, on the other hand, could not be made at that point, as studies on this topic were limited.

Since there is high variability in the results with tDCS, several efforts have been made in literature to identify potential clinical characteristics of participants who respond to tDCS. The influence of gender was documented by Frank et al. (2012), with women having significantly better response to tDCS for their tinnitus suppression. Furthermore, Vanneste et al. (2011b) showed that transcranial magnetic stimulation (TMS) may predict therapy outcomes with tDCS. Responders to tDCS also differ in the resting-state brain activity in the right auditory cortex and parahippocampal area, as well as in the functional connectivity between DLPFC and the subgenual ACC and parahippocampal area, as discussed by Vanneste et al. (2011a). Nevertheless, these factors are not sufficient to explain the high inter-individual variability in response to tDCS. Moreover, replication of these results is necessary, as most studies’ sample sizes are too small to find significant predictive factors.

The broad stimulation pattern, which is one of the main limitations of tDCS (Parazzini et al., 2012), can be overcome with the recently developed technique called High-Definition tDCS (HD-tDCS), using multiple microelectrodes instead of large sponge electrodes. As a result, the focality of HD-tDCS is increased with limited depth of penetration (Datta et al., 2009, Kuo et al., 2013, Villamar et al., 2013a) and a more unidirectional modulation (Edwards et al., 2013, Villamar et al., 2013b). In a pilot study on HD-tDCS, it was shown that the results with this technique were similar to the results with tDCS, reaching a clinically significant improvement in 31% of the 39 patients on the Tinnitus Functional Index (TFI) (Jacquemin et al., 2018). Furthermore, HD-tDCS had some practical advantages compared to tDCS for the clinician and the patients (i.e. fewer side-effects and easier to administer the stimulation of the targeted area) (Jacquemin et al., 2018). This study confirmed the positive results of HD-tDCS indicated by Shekhawat et al. (2016), showing a reduction in tinnitus loudness or annoyance of at least one point in 78% of the 27 patients. That effect was also significant when compared with a sham session (Shekhawat and Vanneste, 2017). Moreover, these authors indicated the safety of this electrical stimulation, with tingling, scalp pain and sleepiness being the most common transient sensations experienced during the stimulation. Hence, the results with HD-tDCS are promising. Yet, there is a need for studies with larger sample sizes to confirm these results and detect possible associated factors for effect of HD-tDCS on tinnitus perception.

The primary objective of this study was to establish the percentage of responders with a large sample size. The secondary objectives were to investigate which tinnitus parameters were changing after HD-tDCS and if there were factors associated with the change in tinnitus severity. The predictive value of the following factors was analyzed: age, gender, type of tinnitus, side of tinnitus, tinnitus etiology and/or tinnitus duration.

## 2. Material and methods

### 2.1 Study design

This prospective longitudinal cohort study was conducted at the Antwerp University Hospital. The clinical effects of six sessions of HD-tDCS with two sessions weekly were investigated. A set of standardized tinnitus questionnaires were filled out at three time points: pre-therapy (T_pre_), post-therapy (T_3w_) and follow-up visit (T_10w_; i.e. seven weeks post-stimulation). Tinnitus handicap, tinnitus severity, tinnitus loudness, the presence of hyperacusis and anxiety or depression disorders were evaluated respectively by means of the Dutch versions of the TFI, Tinnitus Questionnaire (TQ), VAS, Hyperacusis Questionnaire (HQ) and Hospital Anxiety and Depression Scale (HADS). The questionnaires were filled out on a touch-screen desktop and the scoring was computerized.

Besides collecting the questionnaire data, the perceived effect (i.e. self-report) was also documented at T_10w_ by asking an open-ended question: “To which extent did you perceive a change in your tinnitus perception during and after the HD-tDCS treatment?”.

### 2.2 Subjects

A total of 117 patients with chronic, subjective, non-pulsatile tinnitus met the criteria for eligibility (i.e. taking into account contraindications, such as severe brain injuries, presence of metallic devices or implants in the head, or significant skin lesions (Villamar et al., 2013a)) and were included in the present study. Patients with a middle ear pathology or patients who had another tinnitus treatment ongoing were excluded. The demographics and tinnitus-related details of the patients are summarized in Table 1. The hearing thresholds are illustrated in Figure 1.

**Table 1:**
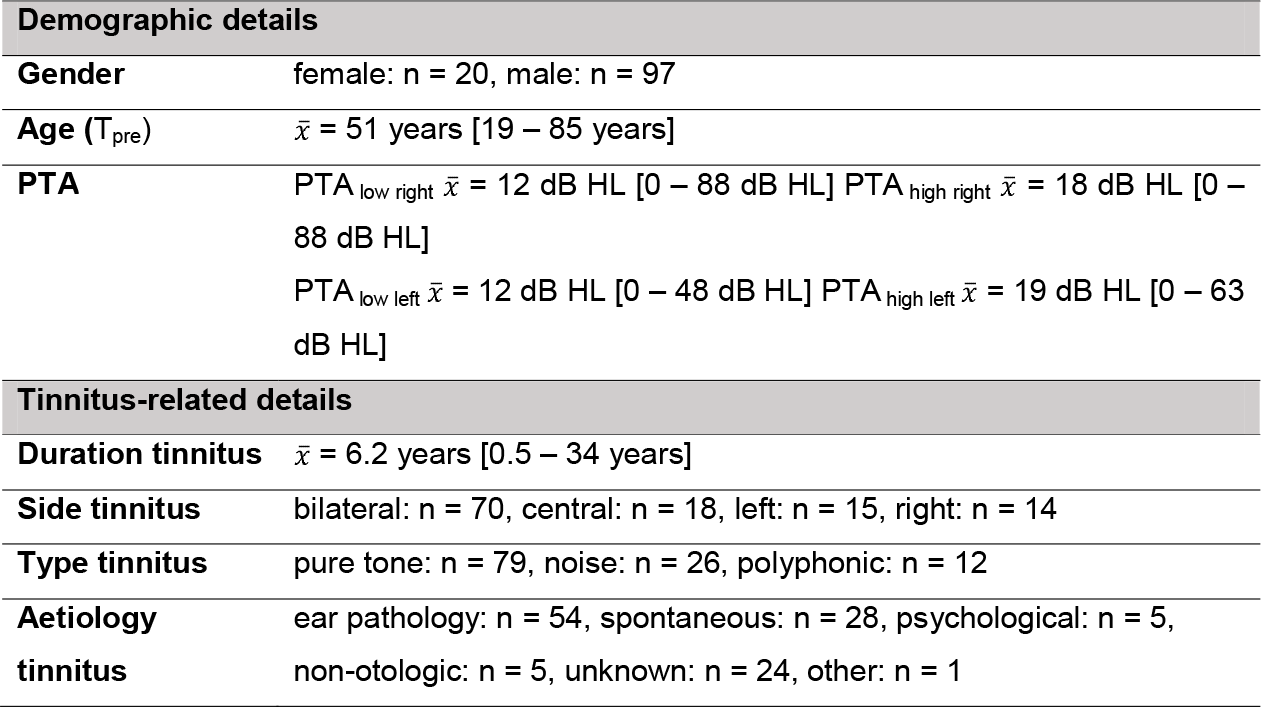
Demographic and tinnitus-related details of the 117 chronic tinnitus patients (PTA, pure tone average; PTA_low_, PTA at 0.5-1-2 kHz; PTA_high_, PTA at 1-2-4 kHz).

**Figure 1:**
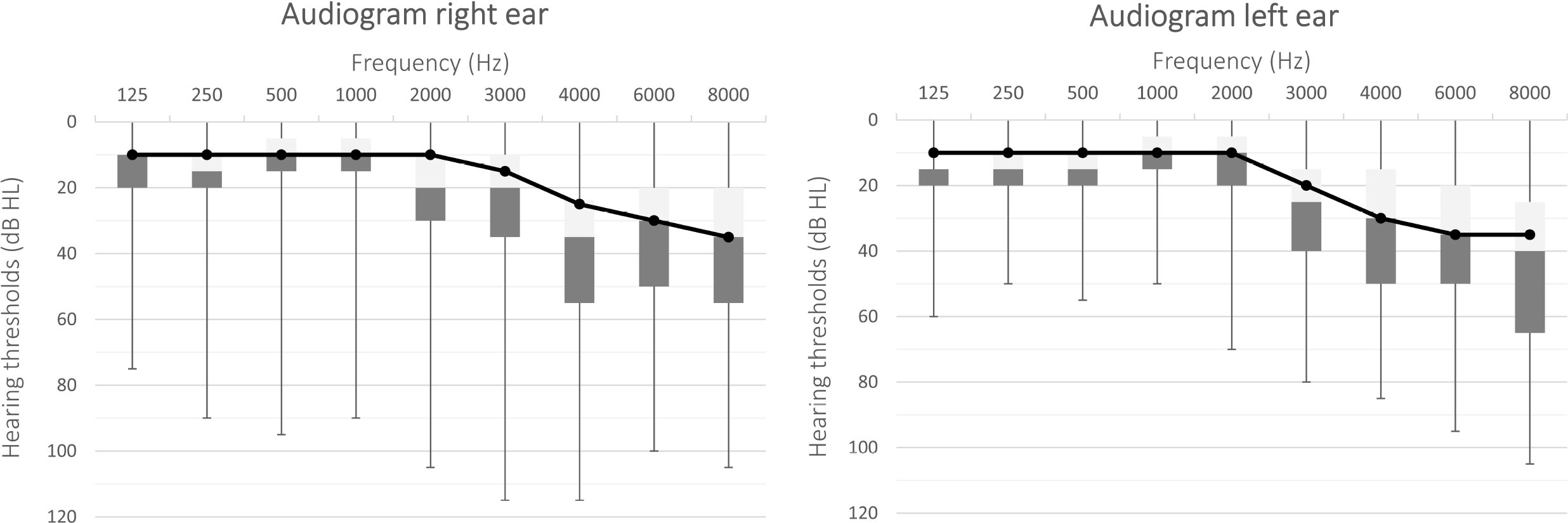
Boxplot (min,Q1,Q3,max) of the air conduction thresholds for the right and left ear of the 117 patients. The black solid line represents the median air conduction threshold for each frequency. (dB HL, dB Hearing Level).

### 2.3 HD-tDCS

Each patient received a total of six sessions of anodal HD-tDCS of right DLPFC with two sessions weekly and a minimum washout period of one day. This schedule was based on the recommendations of Shekhawat and Vanneste (2018), suggesting six tDCS sessions over three weeks’ time. The electrodes were positioned according to the 10/20 international system for EEG electrode placement, with the central anode at F4 and the adjoining cathodes at AF4, FC4, F6 and F2 (Figure 2). The electrodes were silver/silver chloride (Ag/AgCl) ring electrodes with a respectively inner and outer radius of 6 and 12 mm. The current intensity of 2 mA was applied with a fade-in and fade-out of 20 seconds for 20 minutes, delivered by a battery-driven 1×1 tDCS low-intensity stimulator and 4×1 multichannel stimulation adaptor (Soterix Medical Inc, New York, NY), following the HD-tDCS stimulation guidelines (Villamar et al., 2013a).

**Figure 2:**
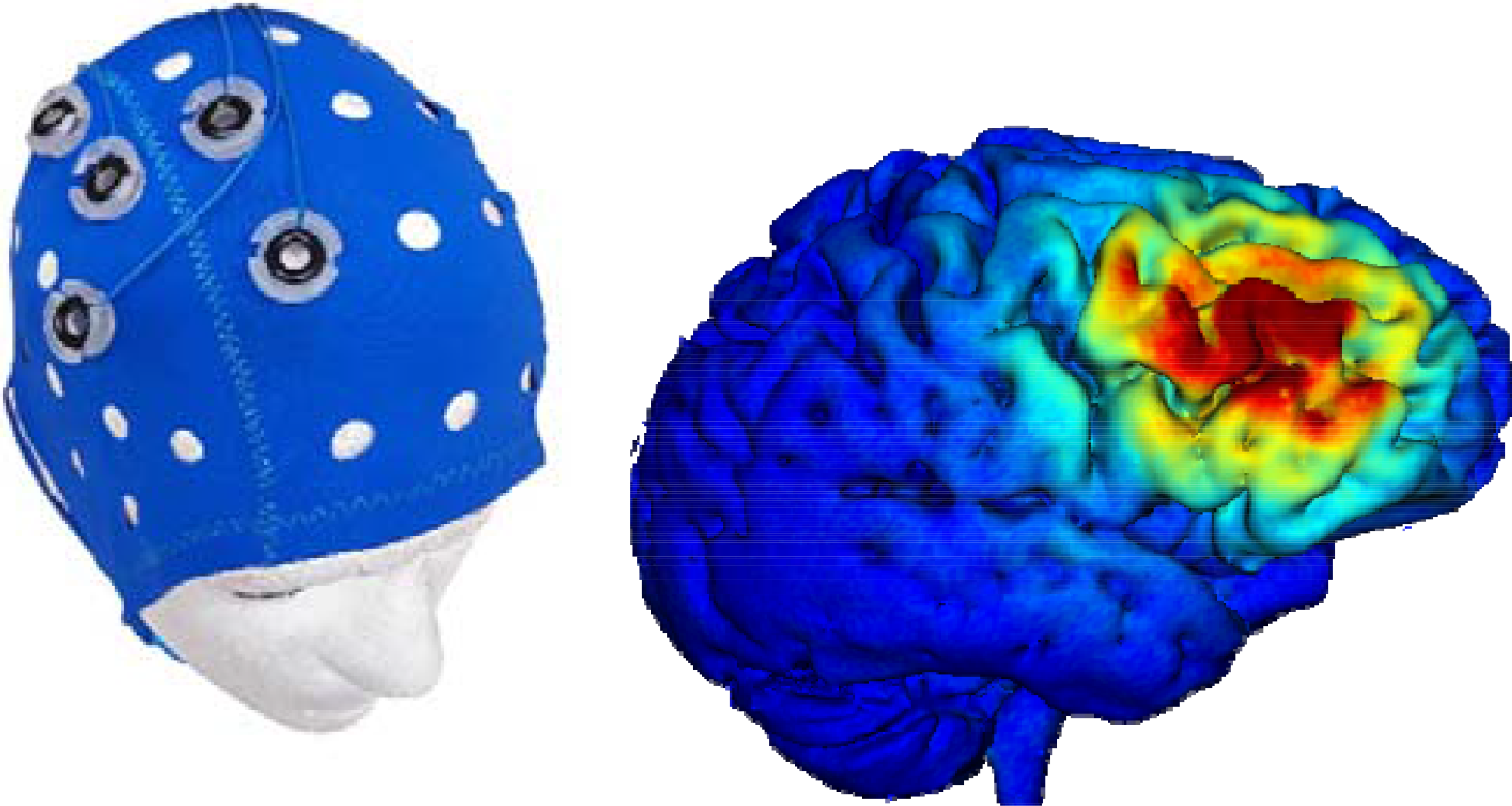
HD-tDCS set-up Electrode positioning at the right DLPFC and a simulation of the current flow of anodal HD-tDCS at the right DLPFC with Soterix HD-Explor TM 4. © Soterix Medical Inc. (HD-tDCS, high-definition transcranial direct current stimulation; DLPFC, dorsolateral prefrontal cortex).

### 2.4 Outcome measurements

The TFI consists of 25 questions evaluating the severity and negative impact of tinnitus, covering eight tinnitus domains (i.e. intrusiveness, sense of control, cognitive complaints, sleep disturbance, auditory difficulties, relaxation, quality of life and emotional distress) (Meikle et al., 2012, Rabau et al., 2014). Answers have to be given on a Likert scale from 0 to 10, resulting in a total score ranging from 0-100, with higher scores denoting higher levels of tinnitus severity. It has been indicated that an improvement of 13 points or more can be interpreted as a clinically significant improvement (Meikle et al., 2012). Assessing treatment-related changes in tinnitus is the main goal of the TFI (Meikle et al., 2012, Fackrell and Hoare, 2014, Jacquemin et al., 2019).

The TQ is a 52-item questionnaire scoring the tinnitus severity, covering five tinnitus domains (i.e. cognitive and emotional distress, intrusiveness, auditory difficulties, sleep disturbance, and somatic complaints). Patients have to score each question on a three-point scale, resulting in a total score ranging from 0-84, with higher scores indicating higher distress levels. A clinically significant improvement has been suggested when TQ improves with 12 points or more (Hall et al., 2018). Assessing changes and relationships between different aspects of complaint and other psychological variables to tinnitus is the main goal of the TQ (Fackrell and Hoare, 2014, Hallam, 2008, Meeus et al., 2007).

The mean tinnitus loudness was scored through a VAS from 0-100 with the help of a ruler and anchored at 0 - ‘no audible tinnitus’ and 100 - ‘extremely loud’ (Adamchic et al., 2012). When the tinnitus was bilateral, the maximum score of both ears had been taken into account in the statistical analysis of the current study.

Hypersensitivity to sounds was rated with the HQ, which consists of 14 self-rating items. Answers had to be given on a 4-point scale, with a total score of more than 28 representing strong auditory hypersensitivity (Khalfa et al., 2002).

The HADS is a self-report screening scale for anxiety and depression disorders, consisting of 14 items with four answer possibilities for each question. A cutoff score of eight points or more has been suggested to screen for these disorders (Spinhoven et al., 1997, Wilkinson and Barczak, 1988, Zigmond and Snaith, 1983).

### 2.5 Statistical analysis

The current study aimed to investigate (1) the effect of HD-tDCS on tinnitus distress as measured by a set of tinnitus questionnaires and (2) predictive factors for evolutions in these questionnaires’ outcomes. There were three repeated measurements for each individual, with the exception of data missing for one individual at T_3w_ and for eight individuals at T_10w_. The first research question was investigated using linear mixed models, which included time as fixed effect and a random intercept, accounting for the nonindependence between the observations taken from the same individual. Either TFI, TQ, VAS, HADS or HQ were entered as dependent variable. If the effect of time was significant, a post hoc analysis with Bonferroni correction for multiple testing was conducted. Responders were determined by calculating the change in questionnaire (i.e. T_pre_ - T_10w_). To test the second research question, a similar linear mixed model was constructed, starting with a model including all fixed effects and their interactions for possible predictive factors, which was simplified using stepwise backward elimination with a significance level of p ≤ .05.

### 2.6 Ethics committee approval

The Committee for Medical Ethics of the University Hospital Antwerp approved the study (B300201630084). All participants gave written informed consent prior to any treatment.

## 3. Results

### 3.1 Questionnaires

The six sessions of anodal HD-tDCS of the right DLPFC resulted in a significant improvement on the TFI total score (p < .01) and the TQ total score (p < .01). As illustrated in figure 3, there was high variability in the evolution of these scores from T_pre_ to T_10w_. The TFI indicated an improvement (i.e. a decrease in total score) in 58% of the patients with a clinically significant improvement in 27%, while the TQ indicated an improvement (i.e. a decrease in total score) in 53% with a clinically significant improvement in 17%. Post-hoc comparisons showed significant changes from T_pre_ to T_10w_ (p_TFI_ < .05; p_TQ_ < .05) and from T_3w_ to T_10w_ (p_TFI_ < .01; p_TQ_ < .01). The other questionnaires (i.e. HADS, HQ, VAS) did not change significantly.

**Figure 3:**
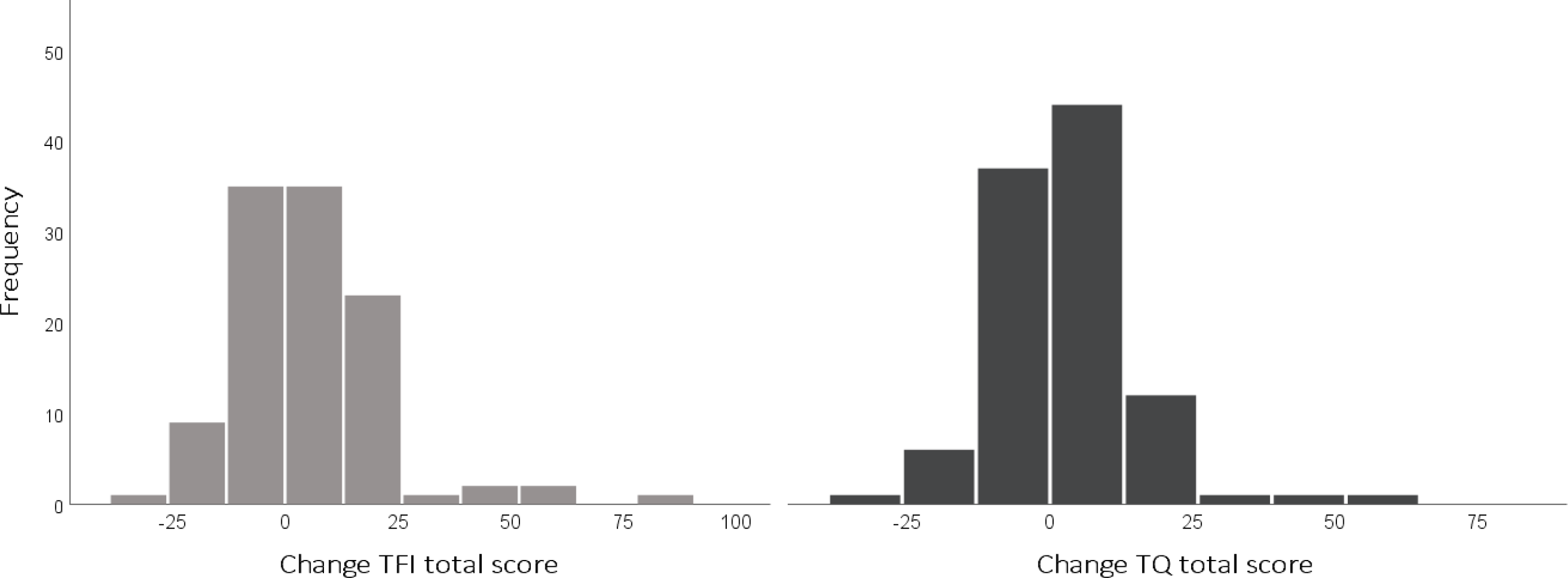
Evolution of TFI and TQ total scores from pre-therapy to follow-up visit Histograms representing change over time (i.e. change = Tpre - T10w). (TFI, tinnitus functional index; TQ, tinnitus questionnaire).

To explore these significant changes in TFI and TQ in-depth, we investigated the effect on the corresponding subscales. The following TFI subscales changed significantly over time: intrusiveness (p < .01), sense of control (p < .01), cognition (p < .05), relaxation (p < .05) and emotional distress (p < .05). For the TQ, intrusiveness (p < .01), emotional and cognitive distress (p < .05) also changed significantly.

### 3.2 Self-report

Participants were asked about side-effects and perceived improvement at T_10w_. A total of 47% reported an improvement after HD-tDCS, for example a decrease in mean and/or maximum loudness of the tinnitus or a better coping mechanism. In general, HD-tDCS was well tolerated by the patients. A total of five out of the 117 patients reported once side-effects during the stimulation, such as tingling, itching, headache, burning or feeling blurry. However, two patients reported headache complaints for a few hours after each stimulation session and one patient experienced an aggravation of the tinnitus symptoms during the treatment and thus needed counseling afterward. The stimulation did not have to be adjusted, as the side-effects were tolerable.

### 3.3 Predictive factors

To find possible predictive factors, a model was constructed based on time (i.e. three repeated measurements), age, gender, type of tinnitus, side of tinnitus, tinnitus etiology and tinnitus duration. After correction for different intercepts between male and female patients, the effect of gender had a significant influence on the evolution of the TFI and TQ over time (p_TFI_ < .01; p_TQ_ < .05), including the following TFI subscales: intrusiveness (p < .01), quality of life (p < .01) and emotional distress (p < .01) and the following TQ subscale: emotional and cognitive distress subscale (p < .01). More specifically, the female patients showed a larger improvement on the TFI after HD-tDCS (figure 4). The remaining factors were removed from the model due to non-significance.

**Figure 4:**
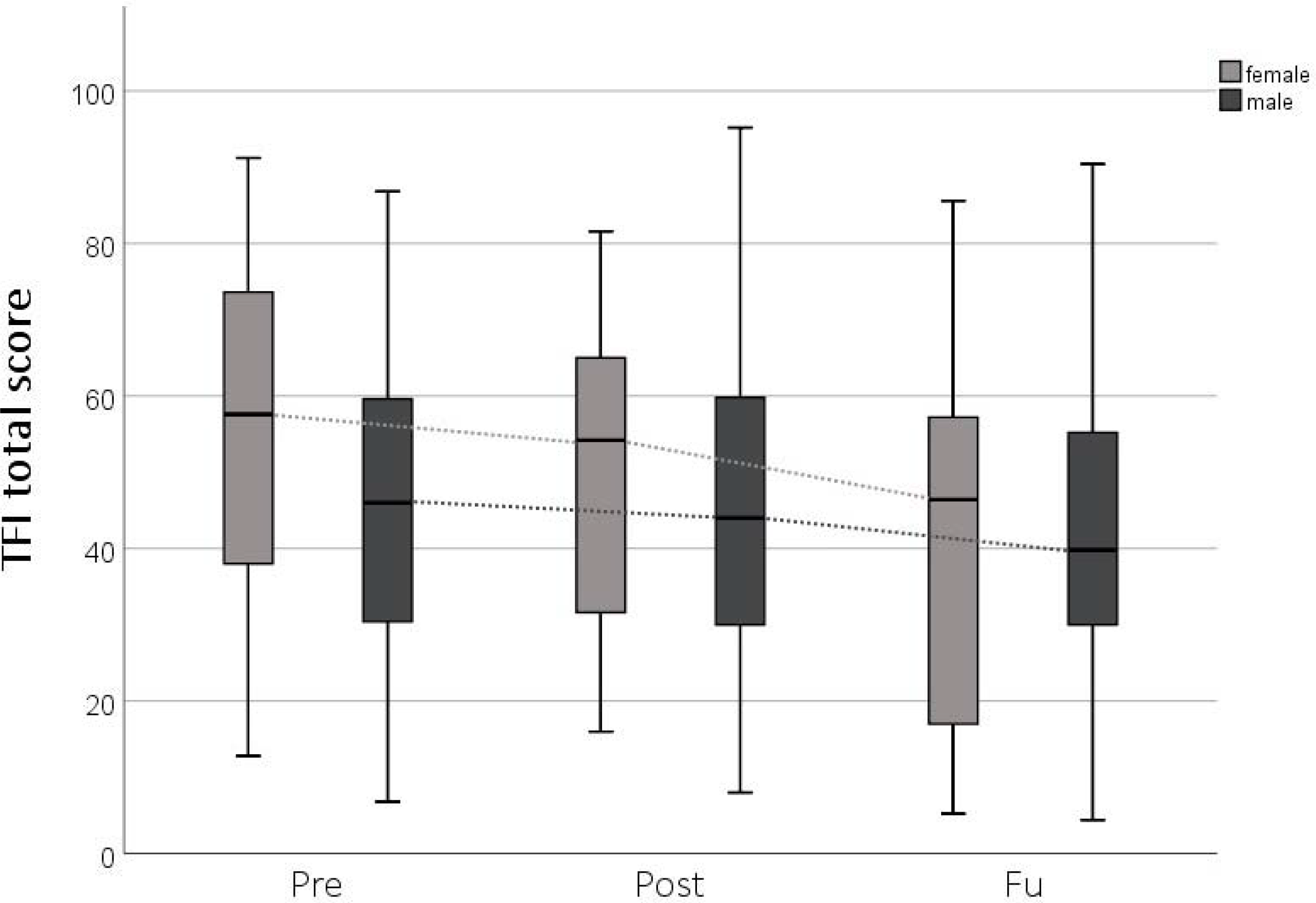
Gender difference of the evolution of the TFI total score over time Boxplots (min, Q1, median, Q3, max). (TFI, tinnitus functional index).

## 4. Discussion

To our knowledge, this is the first study that investigated repeated sessions of HD-tDCS in a large sample of chronic tinnitus patients. A total of 47% of the tinnitus patients in the current study reported a positive effect of anodal HD-tDCS of the right DLPFC at the follow-up visit. Furthermore, the total scores of the TFI and TQ changed significantly over time, in particular from pre-therapy to follow-up visit and from post-therapy to follow-up visit. The analysis of the corresponding subscales showed a decrease in the intrusiveness of the tinnitus, cognitive difficulties, emotional and cognitive distress and an increase in the ability to relax and sense of control over the tinnitus. Moreover, the exploration of possible predictive factors showed a remarkable influence of gender on the HD-tDCS effect with women experiencing a larger improvement after HD-tDCS.

There is high inter-individual variability in the effects of HD-tDCS, which also has been reported in other studies investigating neuromodulation techniques (Frank et al., 2012, Langguth et al., 2008, Rabau et al., 2017). This variability impedes the predictability of the clinical effects for an individual in particular. Hence, this emphasizes the importance of taking into account the tinnitus heterogeneity (Cederroth et al., 2019) and the determination of different ‘tinnitus profiles’ (Van de Heyning et al., 2015). The gender difference, which was also found in a tDCS study by Frank et al. (2012), may play an important role in this context. Firstly, it has been shown that women report higher levels of tinnitus severity or emotional reaction to their tinnitus (Dineen et al., 1997, Erlandsson and Holgers, 2001) prior to therapy, which was also found in the current study. Secondly, the neural activity in tinnitus-related areas, such as the ACC and the orbitofrontal cortex (OFC), differs between men and women during emotional processing and regulation (Butler et al., 2005, Mak et al., 2009, De Ridder et al., 2014). Moreover, Vanneste et al. (2012) showed that the beta activity in the prefrontal cortex in female tinnitus patients is increased, with increased functional alpha connectivity between the OFC, insula, subgenual ACC, parahippocampal areas and the auditory cortex. These areas may be modulated by neuromodulation of the DLPFC (Keeser et al., 2011, Vanneste and De Ridder, 2011). Finally, previous research also showed that effects of tDCS over various brain areas may be gender-dependent in healthy subjects (Chaieb et al., 2008, Boggio et al., 2008, Fumagalli et al., 2010). Overall, the influence of gender found in the current study might be explained by a difference in neural activity in tinnitus-related areas between men and women, which leads to various effects of transcranial stimulation of these areas.

Besides analyzing predictive factors, the aim of this study was to establish the percentage of responders with a large sample size. These results showed a clinically significant improvement on the TFI in 27% of the patients, whereas a previous study with 39 participants showed a slightly larger effect in 31% (Jacquemin et al., 2018). Particularly, the current study indicated a decrease in the intrusiveness of the tinnitus, the emotional distress and the cognitive difficulties and an increase in the ability to relax and the sense of control over the tinnitus. A previous study found similar results, however quality of life was also changing and intrusiveness and relaxation were not improving (Jacquemin et al., 2018). Moreover, the TQ did evolve significantly over time in the current study, whereas a previous study did not find a significant effect for the TQ (Jacquemin et al., 2018). In contrast to the study of Shekhawat and Vanneste (2017), the tinnitus loudness did not change significantly, possibly due to a different assessment of the tinnitus loudness. However, the current results have to be interpreted with caution, as this study was a clinical trial without a sham-group. Previous research on (HD-) tDCS included sham-controlled trials. It was shown that there were no significant effects on loudness, annoyance, anxiety or depression after sham tDCS. In addition, fewer longer-lasting effects were present (Shekhawat and Vanneste, 2017, Garin et al., 2011, Faber et al., 2012, Song et al., 2012a). Furthermore, a delayed-start group of cervical physical therapy for tinnitus did not improve on the TFI total score during a 6 weeks wait-and-see period (Michiels et al., 2016), indicating no spontaneous improvement of the tinnitus perception. Taken together, these results suggest that a sham group or waiting group does not result in effects on the tinnitus perception. Nevertheless, studies following-up on this trial should aim to exclude the placebo effect.

As HD-tDCS appears to be a safe and well-tolerated neuromodulation technique, this technique is promising for tinnitus research. Besides the limited side-effects for patients, the treatment is also easy to administer because of the 10/20 EEG cap (Jacquemin et al., 2018). However, the discrepancy between the self-report (i.e. improvement in 47%) and the subjective questionnaires (i.e. clinically significant improvement in 17-27%) questions the appropriate outcome measure, which is especially important for future explorative studies of innovative treatments. Moreover, the assessment of the therapy outcome during a follow-up visit seems important, as the treatment-related changes were most salient at that point in time and the influence of the act of undergoing therapy could be less. The long-term effect of the HD-tDCS stimulation in the current study is in contrast with the results of previous tDCS studies in tinnitus (Jacquemin et al., 2018, Shekhawat et al., 2015), suggesting that this could be a beneficial effect of HD-tDCS (Kuo et al., 2013). However, it does not rule out the possible influence of other differences in study design, such as sample size and outcome measures.

In the last years, tinnitus research is aiming to improve the effectiveness of tDCS. The evolution to HD-tDCS resulted in practical advantages, but without reaching significantly better results compared to tDCS (Jacquemin et al., 2018). Future studies should elaborate on the predictive factors for HD-tDCS success. Moreover, the effects of simultaneous dual-site stimulation may be explored.

## 5. Conclusions

The current study is the first to report on the effects of HD-tDCS in a large tinnitus population. A total of 47% of the tinnitus patients reported a positive effect of anodal HD-tDCS of the right DLPFC at the follow-up visit, with a larger improvement on the TFI total score for the female tinnitus patients. Future research should include a sham group and focus on increasing the effectiveness of HD-tDCS and searching for additional predictive factors.

## Data Availability

Data can be made available on request

## Acknowledgments

We gratefully acknowledge the statistical support of Kristien Wouters.

## Funding

The present research is financially supported by VLAIO (Agentschap Innoveren en Ondernemen) and a research grant from the FWO (Fonds voor Wetenschappelijk onderzoek Vlaanderen, Egmontstraat 5, 1000 Brussels) (T001916 N).

## Notes

### Competing Interest Statement

The authors have declared no competing interest.

### Clinical Trial

NCT04565132

## References

Adamchic, I., Langguth, B., Hauptmann, C. & Tass, P. A. 2012. Psychometric evaluation of visual analog scale for the assessment of chronic tinnitus. Am J Audiol, 21, 215–25.

Axelsson, A. & Ringdahl, A. 1989. Tinnitus—a study of its prevalence and characteristics. British Journal of Audiology, 23, 53–62.

Baguley, D., Mcferran, D. & Hall, D. 2013. Tinnitus. Lancet, 382, 1600–7.

Boggio, P. S., Rocha, R. R., Da Silva, M. T. & Fregni, F. 2008. Differential modulatory effects of transcranial direct current stimulation on a facial expression go-no-go task in males and females. Neurosci Lett, 447, 101–5.

Butler, T., Pan, H., Epstein, J., Protopopescu, X., Tuescher, O., Goldstein, M., Cloitre, M., Yang, Y., Phelps, E., Gorman, J., Ledoux, J., Stern, E. & Silbersweig, D. 2005. Fear-related activity in subgenual anterior cingulate differs between men and women. Neuroreport, 16, 1233–6.

Cardon, E., Jacquemin, L., Mertens, G., Van De Heyning, P., Vanderveken, O. M., Topsakal, V., De Hertogh, W., Michiels, S., Van Rompaey, V. & Gilles, A. 2019. Cognitive Performance in Chronic Tinnitus Patients: A Cross-Sectional Study Using the RBANS-H. Otology & Neurotology, 40, e876–e882.

Cederroth, C. R., Gallus, S., Hall, D. A., Kleinjung, T., Langguth, B., Maruotti, A., Meyer, M., Norena, A., Probst, T., Pryss, R., Searchfield, G., Shekhawat, G., Spiliopoulou, M., Vanneste, S. & Schlee, W. 2019. Editorial: Towards an Understanding of Tinnitus Heterogeneity. Front Aging Neurosci, 11, 53.

Chaieb, L., Antal, A. & Paulus, W. 2008. Gender-specific modulation of short-term neuroplasticity in the visual cortex induced by transcranial direct current stimulation. Vis Neurosci, 25, 77–81.

Datta, A., Bansal, V., Diaz, J., Patel, J., Reato, D. & Bikson, M. 2009. Gyri-precise head model of transcranial direct current stimulation: improved spatial focality using a ring electrode versus conventional rectangular pad. Brain Stimul, 2, 201-7, 207.e1.

De Ridder, D., Vanneste, S., Weisz, N., Londero, A., Schlee, W., Elgoyhen, A. B. & Langguth, B. 2014. An integrative model of auditory phantom perception: tinnitus as a unified percept of interacting separable subnetworks. Neurosci Biobehav Rev, 44, 16–32.

Dineen, R., Doyle, J. & Bench, J. 1997. Audiological and Psychological Characteristics of a Group of Tinnitus Sufferers, Prior to Tinnitus Management Training. British Journal of Audiology, 31, 27–38.

Edwards, D., Cortes, M., Datta, A., Minhas, P., Wassermann, E. M. & Bikson, M. 2013. Physiological and modeling evidence for focal transcranial electrical brain stimulation in humans: a basis for high-definition tDCS. Neuroimage, 74, 266–75.

Erlandsson, S. & Holgers, K.-M. 2001. The impact of perceived tinnitus severity on health-related quality of life with aspects of gender. Noise and Health, 3, 39–51.

Faber, M., Vanneste, S., Fregni, F. & De Ridder, D. 2012. Top down prefrontal affective modulation of tinnitus with multiple sessions of tDCS of dorsolateral prefrontal cortex. Brain Stimul, 5, 492–8.

Fackrell, K. & Hoare, D. 2014. Questionnaires to Measure Tinnitus Severity. ENT & Audiology news, 22.

Frank, E., Schecklmann, M., Landgrebe, M., Burger, J., Kreuzer, P., Poeppl, T. B., Kleinjung, T., Hajak, G. & Langguth, B. 2012. Treatment of chronic tinnitus with repeated sessions of prefrontal transcranial direct current stimulation: outcomes from an open-label pilot study. J Neurol, 259, 327–33.

Fregni, F., Marcondes, R., Boggio, P. S., Marcolin, M. A., Rigonatti, S. P., Sanchez, T. G., Nitsche, M. A. & Pascual-Leone, A. 2006. Transient tinnitus suppression induced by repetitive transcranial magnetic stimulation and transcranial direct current stimulation. Eur J Neurol, 13, 996–1001.

Fumagalli, M., Vergari, M., Pasqualetti, P., Marceglia, S., Mameli, F., Ferrucci, R., Mrakic-Sposta, S., Zago, S., Sartori, G., Pravettoni, G., Barbieri, S., Cappa, S. & Priori, A. 2010. Brain switches utilitarian behavior: does gender make the difference? PLoS One, 5, e8865.

Garin, P., Gilain, C., Van Damme, J. P., De Fays, K., Jamart, J., Ossemann, M. & Vandermeeren, Y. 2011. Short- and long-lasting tinnitus relief induced by transcranial direct current stimulation. J Neurol, 258, 1940–8.

Hall, D. A., Mehta, R. L. & Argstatter, H. 2018. Interpreting the Tinnitus Questionnaire (German version): what individual differences are clinically important? Int J Audiol, 57, 553–557.

Hallam, R. S. 2008. TQ Manual of the Tinnitus Questionnaire: Revised and updated, London, Polpresa Press.

Jacquemin, L., Mertens, G., Van De Heyning, P., Vanderveken, O. M., Topsakal, V., De Hertogh, W., Michiels, S., Van Rompaey, V. & Gilles, A. 2019. Sensitivity to change and convergent validity of the Tinnitus Functional Index (TFI) and the Tinnitus Questionnaire (TQ): Clinical and research perspectives. Hearing Research, 382, 107796.

Jacquemin, L., Shekhawat, G. S., Van De Heyning, P., Mertens, G., Fransen, E., Van Rompaey, V., Topsakal, V., Moyaert, J., Beyers, J. & Gilles, A. 2018. Effects of Electrical Stimulation in Tinnitus Patients: Conventional Versus High-Definition tDCS. Neurorehabil Neural Repair, 32, 714–723.

Keeser, D., Padberg, F., Reisinger, E., Pogarell, O., Kirsch, V., Palm, U., Karch, S., Moller, H. J., Nitsche, M. A. & Mulert, C. 2011. Prefrontal direct current stimulation modulates resting EEG and event-related potentials in healthy subjects: a standardized low resolution tomography (sLORETA) study. Neuroimage, 55, 644–57.

Khalfa, S., Dubal, S., Veuillet, E., Perez-Diaz, F., Jouvent, R. & Collet, L. 2002. Psychometric normalization of a hyperacusis questionnaire. ORL J Otorhinolaryngol Relat Spec, 64, 436–42.

Kuo, H. I., Bikson, M., Datta, A., Minhas, P., Paulus, W., Kuo, M. F. & Nitsche, M. A. 2013. Comparing cortical plasticity induced by conventional and high-definition 4 x 1 ring tDCS: a neurophysiological study. Brain Stimul, 6, 644–8.

Langguth, B. & De Ridder, D. 2011. Neuromodulation: Introduction. In: MøLler, A. R., Langguth, B., De Ridder, D. & Kleinjung, T. (eds.) Textbook of Tinnitus. New York, NY: Springer New York.

Langguth, B., De Ridder, D., Dornhoffer, J. L., Eichhammer, P., Folmer, R. L., Frank, E., Fregni, F., Gerloff, C., Khedr, E., Kleinjung, T., Landgrebe, M., Lee, S., Lefaucheur, J. P., Londero, A., Marcondes, R., Moller, A. R., Pascual-Leone, A., Plewnia, C., Rossi, S., Sanchez, T., Sand, P., Schlee, W., Pysch, D., Steffens, T., Van De Heyning, P. & Hajak, G. 2008. Controversy: Does repetitive transcranial magnetic stimulation/transcranial direct current stimulation show efficacy in treating tinnitus patients? Brain Stimul, 1, 192–205.

Lefaucheur, J. P., Antal, A., Ayache, S. S., Benninger, D. H., Brunelin, J., Cogiamanian, F., Cotelli, M., De Ridder, D., Ferrucci, R., Langguth, B., Marangolo, P., Mylius, V., Nitsche, M. A., Padberg, F., Palm, U., Poulet, E., Priori, A., Rossi, S., Schecklmann, M., Vanneste, S., Ziemann, U., Garcia-Larrea, L. & Paulus, W. 2017. Evidence-based guidelines on the therapeutic use of transcranial direct current stimulation (tDCS). Clin Neurophysiol, 128, 56–92.

Mak, A. K., Hu, Z. G., Zhang, J. X., Xiao, Z. & Lee, T. M. 2009. Sex-related differences in neural activity during emotion regulation. Neuropsychologia, 47, 2900–8.

Meeus, O., Blaivie, C. & Van De Heyning, P. 2007. Validation of the Dutch and the French version of the Tinnitus Questionnaire. B-ent, 3 Suppl 7, 11–7.

Meikle, M. B., Henry, J. A., Griest, S. E., Stewart, B. J., Abrams, H. B., Mcardle, R., Myers, P. J., Newman, C. W., Sandridge, S., Turk, D. C., Folmer, R. L., Frederick, E. J., House, J. W., Jacobson, G. P., Kinney, S. E., Martin, W. H., Nagler, S. M., Reich, G. E., Searchfield, G., Sweetow, R. & Vernon, J. A. 2012. The tinnitus functional index: development of a new clinical measure for chronic, intrusive tinnitus. Ear Hear, 33, 153–76.

Michiels, S., Van De Heyning, P., Truijen, S., Hallemans, A. & De Hertogh, W. 2016. Does multi-modal cervical physical therapy improve tinnitus in patients with cervicogenic somatic tinnitus? Man Ther, 26, 125–131.

Parazzini, M., Fiocchi, S. & Ravazzani, P. 2012. Electric field and current density distribution in an anatomical head model during transcranial direct current stimulation for tinnitus treatment. Bioelectromagnetics, 33, 476–87.

Rabau, S., Shekhawat, G. S., Aboseria, M., Griepp, D., Van Rompaey, V., Bikson, M. & Van De Heyning, P. 2017. Comparison of the Long-Term Effect of Positioning the Cathode in tDCS in Tinnitus Patients. Front Aging Neurosci, 9, 217.

Shekhawat, G. S., Stinear, C. M. & Searchfield, G. D. 2013. Transcranial direct current stimulation intensity and duration effects on tinnitus suppression. Neurorehabil Neural Repair, 27, 164–72.

Shekhawat, G. S., Stinear, C. M. & Searchfield, G. D. 2015. Modulation of Perception or Emotion? A Scoping Review of Tinnitus Neuromodulation Using Transcranial Direct Current Stimulation. Neurorehabilitation and Neural Repair, 29, 837–846.

Shekhawat, G. S., Sundram, F., Bikson, M., Truong, D., De Ridder, D., Stinear, C. M., Welch, D. & Searchfield, G. D. 2016. Intensity, Duration, and Location of High-Definition Transcranial Direct Current Stimulation for Tinnitus Relief. Neurorehabil Neural Repair, 30, 349–59.

Shekhawat, G. S. & Vanneste, S. 2017. High-definition transcranial direct current stimulation of the dorsolateral prefrontal cortex for tinnitus modulation: a preliminary trial. J Neural Transm (Vienna).

Shekhawat, G. S. & Vanneste, S. 2018. Optimization of Transcranial Direct Current Stimulation of Dorsolateral Prefrontal Cortex for Tinnitus: A Non-Linear Dose-Response Effect. Scientific Reports, 8, 8311.

Song, J.-J., Vanneste, S., Van De Heyning, P. & De Ridder, D. 2012a. Transcranial Direct Current Stimulation in Tinnitus Patients: A Systemic Review and Meta-Analysis. The Scientific World Journal, 2012, 427941.

Song, J. J., Vanneste, S., Heyning, P. & Ridder, D. 2012b. Transcranial direct current stimulation in tinnitus patients: a systemic review and meta-analysis. Scientific World Journal, 2012.

Spinhoven, P., Ormel, J., Sloekers, P. P., Kempen, G. I., Speckens, A. E. & Van Hemert, A. M. 1997. A validation study of the Hospital Anxiety and Depression Scale (HADS) in different groups of Dutch subjects. Psychol Med, 27, 363–70.

Van De Heyning, P., Gilles, A., Rabau, S. & Van Rompaey, V. 2015. Subjective tinnitus assessment and treatment in clinical practice: the necessity of personalized medicine. Curr Opin Otolaryngol Head Neck Surg, 23, 369–75.

Vanneste, S. & De Ridder, D. 2011. Bifrontal transcranial direct current stimulation modulates tinnitus intensity and tinnitus-distress-related brain activity. Eur J Neurosci, 34, 605–14.

Vanneste, S., Focquaert, F., Van De Heyning, P. & De Ridder, D. 2011a. Different resting state brain activity and functional connectivity in patients who respond and not respond to bifrontal tDCS for tinnitus suppression. Exp Brain Res, 210, 217–27.

Vanneste, S., Joos, K. & De Ridder, D. 2012. Prefrontal cortex based sex differences in tinnitus perception: same tinnitus intensity, same tinnitus distress, different mood. PLoS One, 7, e31182.

Vanneste, S., Langguth, B. & De Ridder, D. 2011b. Do tDCS and TMS influence tinnitus transiently via a direct cortical and indirect somatosensory modulating effect? A combined TMS-tDCS and TENS study. Brain Stimul, 4, 242–52.

Villamar, M. F., Volz, M. S., Bikson, M., Datta, A., Dasilva, A. F. & Fregni, F. 2013a. Technique and considerations in the use of 4×1 ring high-definition transcranial direct current stimulation (HD-tDCS). J Vis Exp, e50309.

Villamar, M. F., Wivatvongvana, P., Patumanond, J., Bikson, M., Truong, D. Q., Datta, A. & Fregni, F. 2013b. Focal modulation of the primary motor cortex in fibromyalgia using 4×1- ring high-definition transcranial direct current stimulation (HD-tDCS): immediate and delayed analgesic effects of cathodal and anodal stimulation. J Pain, 14, 371–83.

Wilkinson, M. J. B. & Barczak, P. 1988. Psychiatric screening in general practice: comparison of the general health questionnaire and the hospital anxiety depression scale. The Journal of the Royal College of General Practitioners, 38, 311–313.

Zigmond, A. S. & Snaith, R. P. 1983. The hospital anxiety and depression scale. Acta Psychiatr Scand, 67.

